# SEIRDQ: A COVID-19 case projection modeling framework using ANN to model quarantine

**DOI:** 10.1101/2021.10.15.21265056

**Authors:** Harish Chandra, Arman Margaryan, Xianwei Meng

## Abstract

We propose and implement a novel approach to model the evolution of COVID-19 pandemic and predict the daily COVID-19 cases (infected, recovered and dead). Our model builds on the classical SEIR-based framework by adding additional compartments to capture recovered, dead and quarantined cases. Quarantine impacts are modeled using an Artificial Neural Network (ANN), leveraging alternative data sources such as the Google mobility reports. Since our model captures the impact of lockdown policies through the quarantine functions we designed, it is able to model and predict future waves of COVID-19 cases. We also benchmark out-of-sample predictions from our model versus those from other popular COVID-19 case projection models.

## 1 Background

COVID-19 is a new strain of coronavirus, SARS-CoV-2, first detected in Wuhan (Hubei), China [1], [2]. Within a few months after discovering it, the number of COVID-19 cases increased exponentially within mainland China and worldwide. Globally, governments introduced critical measures including social distancing and quarantine measures to address the COVID-19 outbreak [3]. As per the World Health Organization situation report published on 25 May 2020, there were more than five million total cases and over 340,000 deaths around the globe [4].

Mathematically modeling the dynamics of COVID-19 evolution is useful to plan effective pandemic spread control strategies. Various models have been proposed to model the spread of COVID-19 pandemic. Giordano et al. provides a good overview of various approaches, including those based on the classical SIR and SEIR frameworks [5]. In Fig.1, we present a summary of various modeling approaches along with their strengths and weaknesses. In this paper, we propose a novel modeling framework that builds on a classical modeling framework such as the SEIR, while taking advantage of real-world data to model quarantine impacts which are otherwise challenging to model using analytical approximations.

**Figure 1:**
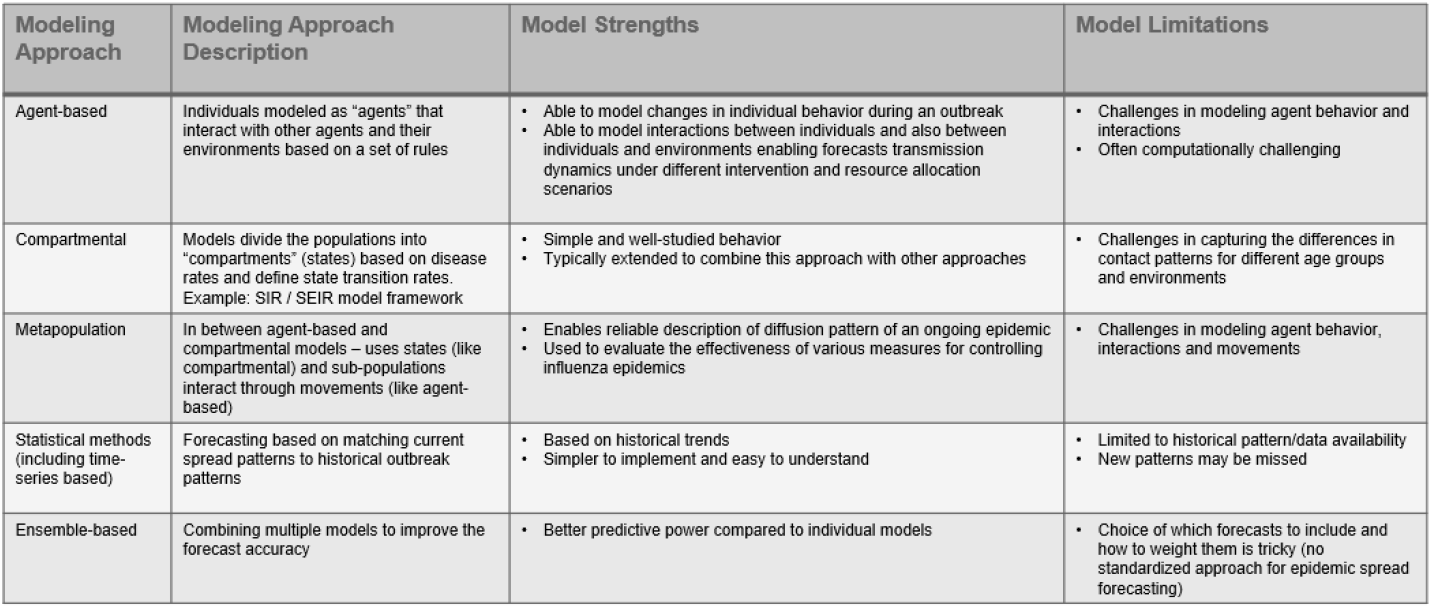
Types of modeling approaches

We rely on publicly available COVID-19 case data (John Hopkins data source) as well as other publicly available alternative sources of data to project the number of infected, recovered and dead cases. The underlying model, based on the classical SEIR-based framework, assumes that the total population in a country or a state can be compartmentalized into mutually exclusive compartments - *S, E, I* and *R* (explained in Section 2.1) and the interactions between these compartments is modelled using a coupled non-linear ODE system of equations. Our model has an Artificial Neural Network (ANN) model component to model the quarantine impacts based on mobility information from Google.

In section 2, we discuss the classical SEIR model and provide some results when we apply our SEIRDQ model to US, Wuhan (China) and Italy. In section 3, we present additional states to bifurcate the “removed” cases in the classical SEIR model into “dead” and “recovered” cases. We then model the quarantine effects in section 4 using a simple logistic function and review the results. In section 4, we show that the quarantine impacts are better modelled using an *ANN model* trained on mobility data reports from Google. Lastly, in section 5, we assess the SEIRDQ model performance based on out-of-sample tests for weekly projections and also benchmark our model against some of the popular COVID-19 projection models.

## 2 Classical SEIR Model

Several modeling approaches have been developed to describe spread of an epidemic. A general overview of modeling approaches with their pros and cons is given in Fig.1.

The origins of epidemiological compartmental models may be traced back to contributions made by Kermack and McKendrick in 1927 [6]. Since then, the compartmental models have been very popular since they are easy to interpret and can be modified to account for several factors such as existence of more than one strain of the same virus, recovered people with or without immunity to be reinfected and introduction of vaccines during the life of the epidemic. Based on these and the advantages described in Fig.1, we chose to extend the classical SEIR model, a compartmental model.

### 2.1 Theory

As introduced in the previous sections, the classical SEIR model assumes that the total population for a geographical entity (a country, state or even a county), assumed constant over the period of observations and projections, is compartmentalized into mutually exclusive states, Susceptible (*S*), Exposed (*E*), Infected (*I*) and Removed (*R*). These variables are described below:

1. Susceptible (*S*) stands for the number of people that were never infected
2. Exposed (*E*) stands for the number of infected people, but not yet infectious (incubation period)
3. Infected (*I*) stands for the number of infected people that are also infectious
4. Removed (*R*) stands for the number of people that recovered or deceased

The interactions between these four states (or compartments in epidemiological parlance) is assumed to be driven by a system of coupled non-linear ODEs as follows:

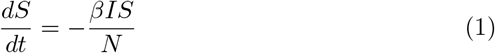

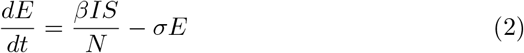

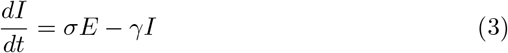

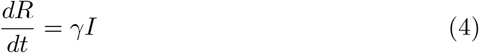

where, *N*, the total population (= *S*(*t*) + *E*(*t*) + *I*(*t*) + *R*(*t*)) is assumed to be constant. The parameters, *β, σ* and *γ* represent the exposure rate, infection rate and recovery rate, respectively. The parameter, *β* is just the product of probability of disease transmission (when a healthy individual meets an infectious individual) and daily average number of people (healthy and otherwise) one infectious person meets. The incubation period is assumed to be a random variable with exponential distribution with the rate parameter, *σ*. Here, the average incubation period is therefore 1*/σ*, and hence, the number of exposed individuals becoming infectious per day is *σE*. Similarly, average time to recovery/death is 1*/γ*, and hence, the number of infectious individuals recovering/dying per day is *γI*. The basic reproduction number, *R*_0_ = *β/γ* is proportional to the average number of individuals one infected individual infects.

There are a few disadvantages associated with the classical SEIR model. Total population of the region it is applied to is assumed constant. Individuals who recovered from COVID-19 are assumed to be immune to reinfection. The model does not distinguish between the people who recovered and the people who died. Instead, it just combines those two compartments into just one compartment - “Removed”, comprised of people that can neither be infected nor can infect others. More importantly, the classical SEIR modeling framework does not take into account the effects of governmental quarantine and self-quarantine policies. Our SEIRDQ model address two of the shortcomings of the classical SEIR model by splitting the “Removed” compartment into “Deceased” and “Recovered”. This allows our model to be more descriptive by predicting more granular compartments while being able to better model the quarantine effects through a data-driven approach (see Section 3).

Note that there is a simpler SIR framework that can be considered a simpler version of the SEIR model with no separate representation for the exposed category of individuals, assuming direct transition from susceptible (*S*) to infected (*I*). Note that these modeling frameworks assume that the reinfection (possibility that individuals who recovered could be reinfected) rate is negligible. In the following section, we present a few results for both SEIR and SIR based models.

### 2.2 Results

In this section, we present some results based on the classical SEIR (and SIR) model implementations applied to the COVID-19 case data from US and Italy.

As can be seen from the results, these classical models do not capture the plateauing trends due to the quarantine policies implemented in the respective countries.

For US and Italy, their COVID-19 data on confirmed cases, deaths and recovered numbers are based on the data collected by John Hopkins University.^1^ When the SEIR (and SIR) models are applied to these two countries’ data, we select the starting date with about 500 confirmed cases. That improves the reliability and performance of the model estimation and prediction, while facilitating better comparison of model results for the two countries. The model training periods for U.S. start on March 08, 2020 with 518 confirmed cases, and for Italy, the start date is February 27 with 655 confirmed cases. Each country used about 50 days’ data for training the model. The SEIR model fit and prediction results for U.S. and Italy are presented in Fig.2 and Fig.3 respectively.

**Figure 2:**
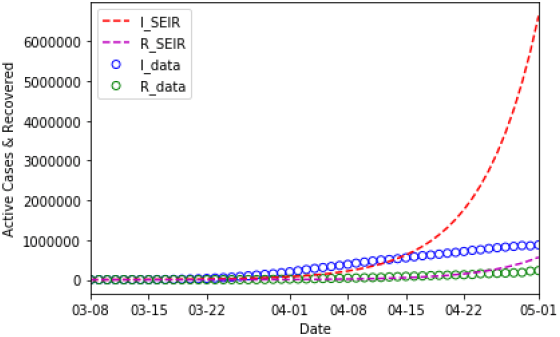
US SEIR model calibration period Mar 08 - Apr 30. SEIR Projection Results Using the Classical SEIR Model

**Figure 3:**
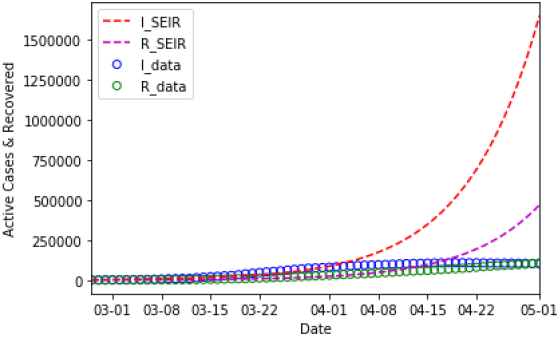
Italy SEIR model calibration period Feb 27 - Apr 16. SEIR Projection Results Using the Classical SEIR Model

Based on the graphs plot in Fig.2, Fig.3, Fig.4 and Fig.5, we see that the classical SEIR models fail to adequately model the real-world data for both U.S. and Italy. Though the SEIR projections fit the observation well at the beginning periods, divergence between their projections and the real-world data blows up later. One reason is that the classical SEIR model does not capture impacts from important drivers of COVID-19 spread such as quarantine and lock down policies (such as social distancing, home shelter and quarantine) during pandemic periods. Such policies are aimed to contain the coronavirus spread. For the U.S., the estimated SEIR parameters are *β* = 0.261, *γ* = 0.013 which imply the basic reproduction number, *R*_0_ = 20.7. For the case of Italy, the estimated SEIR parameters are *β* = 0.200, *γ* = 0.0277 which imply the basic reproduction number, *R*_0_ = 7.2. Both are way too large compared with the widely accepted *R*_0_ value. As noted earlier, reproductive number can be loosely interpreted as the average number of individuals one infected individual infects.

**Figure 4:**
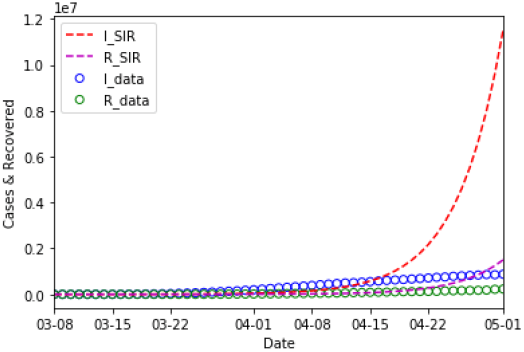
US SIR model calibration period Mar 08 - Apr 30. SIR Projection Results Using the Classical SIR Model

**Figure 5:**
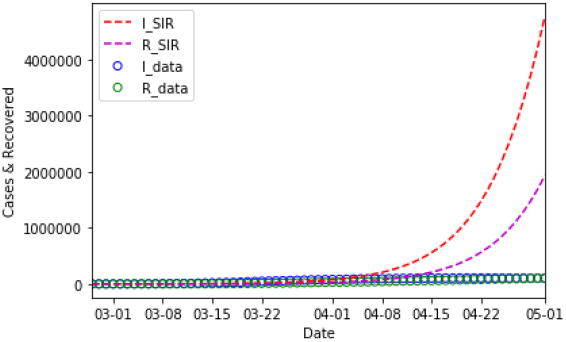
Italy SIR model calibration period Feb 27 - Apr 16. SIR Projection Results Using the Classical SIR Model

For similar reasons cited above, the classical SIR models for U.S. and Italy also fail as illustrated in Fig.4 and Fig.5.

## 3 Modeling Quarantine Impacts

In this section, we demonstrate improvements over the classical SEIR framework by modelling the quarantine impact using a simple logistic function. We first describe the quarantine function design approach.

### 3.1 Quarantine Function

We start with a common S-shaped logistic (or sigmoid) function to model the quarantine strength function *ρ*(*t*).^2^ Logistic function is widely applied to population growth models in ecology (see [15],[16]), regression models in statistics (see [17], [18]), and artificial neural networks (see [20],[19]). As explained in [9], *ρ*(*t*) is inversely proportional to the average time it takes for an infected person to be quarantined and to further infect any susceptible individuals.

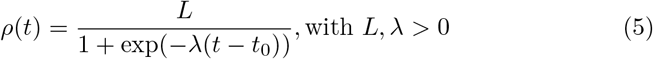

The parameter *L* is the maximum value. *λ* is the growth rate or steepness of the curve and *t*_0_ is the inflection and midpoint of the logistic curve. And the quarantine policy will affect the pandemic infection processes in a simple way through modified infectious equation

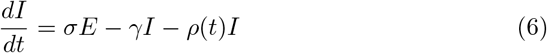

The updated SEIR model incorporated with logistic quarantine function is applied to the Covid-19 cases for U.S. and Italy. The results are shown as in the Fig. 6 and Fig. 7.

**Figure 6:**
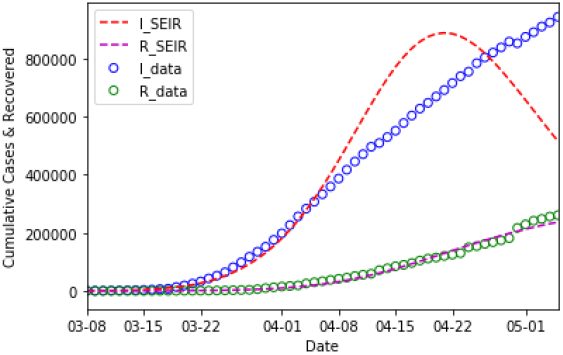
US, model calibration period Mar 08 - Apr 30. SEIR Projection Results Using the Logistic Quarantine Function

**Figure 7:**
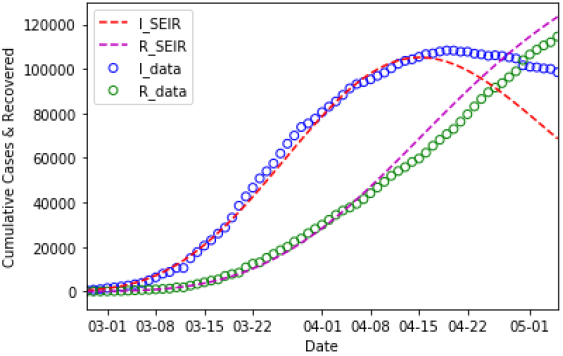
Italy, model calibration period Feb 27 - Apr 16. SEIR Projection Results Using the Logistic Quarantine Function

Based on the results, the SEIR models with the simple logistic quarantine function fit well with the COVID-19 cases and also with the deaths data for the U.S. and Italy. However, the assumption of simple logistic function form for the quarantine policy is arguable if it cannot effectively model the real-world data. We choose alternative data such as the Google mobility index data which may be leveraged to inform our quarantine function. More on Google mobility data is presented in the next section.

### 3.2 Using Alternative Data

The Google mobility index data, which is helpful to infer the social distancing and quarantine impacts, is based on the daily Global Mobility Report (GMR) published online by Google (https://www.google.com/covid19/mobility). The GMR data shows how visits and length of stay at different places compared to a baseline. The baseline is the median value for the corresponding day of the week, during the 5-week period Jan 3rd - Feb 6th, 2020. It provides the movement trends over time by geography, across different categories of places such as retail and recreation, groceries and pharmacies, parks, transit stations, workplaces, and residential.

The data is based on Google users who have opted-in to share location history through their Google accounts, depending on user privacy settings and connectivity. Currently the GMR data covers 136 countries and regions including US national and state level mobility indices. Based on the GMR raw data (Fig.8), various outdoor activities (shopping, office visits, etc.) dropped dramatically while more people started staying at home when the quarantine policy was in effect.

**Figure 8:**
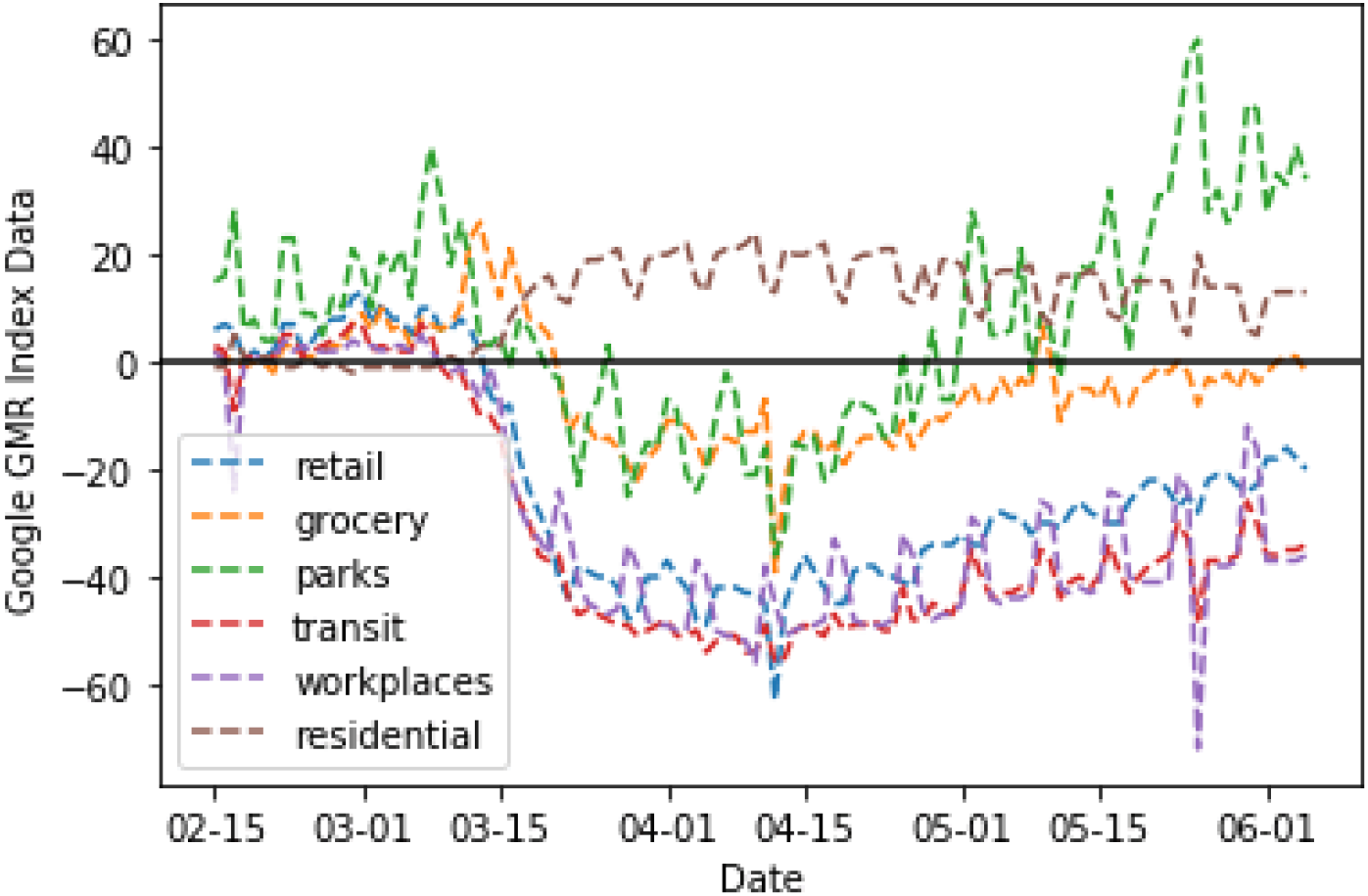
Google Global Mobility Report Data

The quarantine signal, *G*(*t*) is modelled as a product of a logistic function and an annually periodic cosine function as shown below. We assume an annual frequency to capture the long-term quarantine trends.

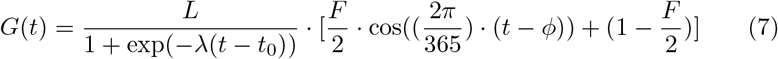

where the amplitude parameter *F* ∈ [0, 2] guarantees positivity and *ϕ* is the phase position parameter. The time *t* is measured in days and the trigonometric function part has a time period of 365 days. The parameter, *t*_0_, denotes the start day. The logistic part captures the rising impacts of the quarantine policy observed from the Google mobility indices during the early stages. Based on the plots of the GMR data, there is an obvious weekly periodic pattern in the mobility behaviors. Therefore, a weekly periodic function is added to better fit the GMR data.

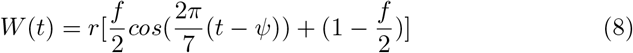

Similar to the annually periodic part in Eq.7, the parameter, *f* ∈ [0, 2], and the overall amplitude multiplier, *r >* 0, apply for the weekly periodic function, *W* (*t*) in Eq.8. Finally, the GMR data is modelled as a linear combination of *G*(*t*) and *W* (*t*). The quarantine signal, *Q*(*t*), is modeled as positive function and we note that quarantine policy will curtail public activities at retail, groceries and pharmacies, parks, transit stations, and workplaces while generally increase the time people stay at home (i.e., residential). Accordingly, the residential GMR data is modeled as the sum of the quarantine signal, *G*(*t*), and weekly signal, *W* (*t*), and other GMR data are modeled as the difference between the two signals.

### 3.3 Results

The GMR model is applied to the retail and transit Google mobility data to calibrate the model parameters. The raw Google GMR retail and transit data, the identified long-term quarantine function, and the fitted curves for the retail and transit cases are plotted in Fig.9 and Fig.10, respectively.

**Figure 9:**
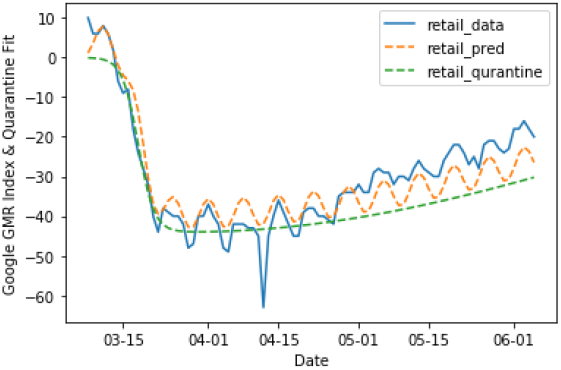
GMR Retail. Identified Quarantine Signal based on Google GMR data

**Figure 10:**
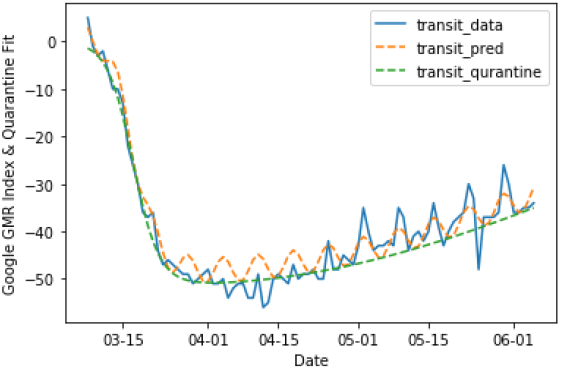
GMR Transit. Identified Quarantine Signal based on Google GMR data

Alternative data sources such as the Google GMR data is valuable to quantify the quarantine policy and applied to the Covid-19 spread modeling. Consistent with the timing of the social distancing and quarantine polices, the identified long-term quarantine functions (green curves in Fig.9 and Fig.10) increase rapidly starting the second week of March 2020. That’s the main driving force for the rapid decline in the activities and people flows in retail (Fig.9), and transit stations (Fig.10). We also observe that the long-term quarantine trend slowly decrease after the mid-April, which is consistent with the rising trend in Google retail and transit data. The combined long-term quarantine trend and weekly patterns provides us the final quarantine signals (plotted orange curves), which fit well with the Google GMR data. Our quarantine functions reasonably captures the long-term trend and weekly patterns of quarantine impacts observed in the Google GMR data.

## 4 SEIRDQ Model

Effectiveness of social distancing, quarantine policies and individual reactions is challenging to model. Compartmental models such as the DELPHI [10] assume an analytical approximation to capture quarantine effects. Another compartmental model, SIDARTHE [5], distinguishes between infected individuals depending on whether they have been diagnosed and on the severity of their symptoms. In this section, we present our *SEIRDQ model*, an extension of the classical SEIR model (described in Section 2) by including two distinct states for dead and recovered cases (versus a single “removed” state in the case of classical SEIR model) and also a quarantine state. In this model, quarantine is modelled using ANN-trained Google mobility reports. Diagrammatic representation of the SEIRDQ model is given in Fig.11. We note that additional states (i.e., in addition to the classical SEIR framework) does not necessarily translate to higher accuracy but often helps with more granular case reporting.

**Figure 11:**
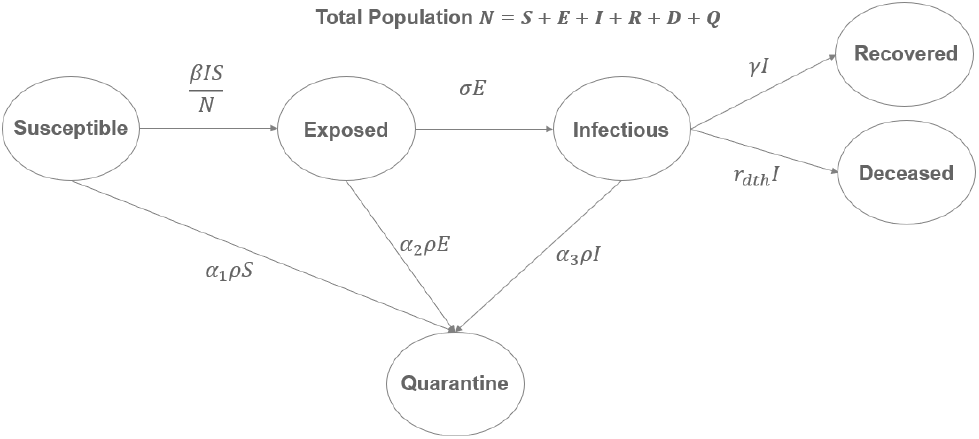
Diagrammatic representation of SEIRDQ model

The interactions between these six states (i.e., *S, E, I, R, D* and *Q*) is assumed to be driven by a system of coupled non-linear ODEs as follows:

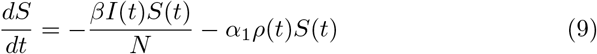

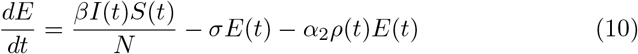

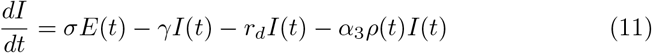

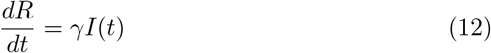

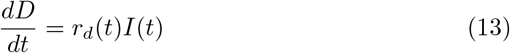

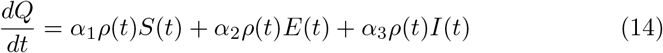

where, *N*, the total population (= *S*(*t*) + *E*(*t*) + *I*(*t*) + *R*(*t*) + *D*(*t*) + *Q*(*t*)) is assumed to be constant. The parameters, *β, σ* and *γ* are the same as what they were for the classical SEIR model described in section 2. The *α* parameters introduced here help us calibrate the dynamics of interactions between the *S, E* and *I* states and the quarantine state, *Q*. Although the major driver is the infected state, *I*, our model captures the interactions between *S* and *E* states and the quarantine state, *Q*. Here we assume that the strengths of quarantine policies exercised by the people in compartments *S, E* and *I* are proportional to each other and that those proportionality coefficients are constant in time. Based on this assumption, we can see from the above equations that the speeds of flow of people from compartments *S, E* and *I* into the compartment *Q* (*α*_1_*ρ*(*t*)*S*(*t*), *α*_2_*ρ*(*t*)*S*(*t*) and *α*_3_*ρ*(*t*)*S*(*t*), respectively) are proportional to each other and that the proportionality coefficients *α*_1_, *α*_2_ and *α*_3_ are assumed to be constant in time.

### 4.1 Modeling *ρ*(*t*) Using ANN

In the previous section 3.1, we presented a quarantine model based on a simple logistic function. In this section, we model the quarantine function, *ρ* using an *ANN model* :

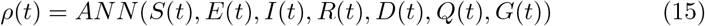

Here, the input layer is comprised of *S, E, I, R, D, Q* and *G*. Note that *G*(*t*) is the quarantine signal from Google mobility reports and is a direct input to the ANN model unlike other inputs that are generated by integrating the ODEs described in equations in section 2.1. The neural network for the quarantine function *ρ*(*t*) has architecture 7 × 14 × 1 with an input layer of seven neurons, one hidden layer with fourteen neurons and an output layer of single neuron. The number of trainable parameters of the ANN (the weights and biases) is 127. The SEIRDQ model as a whole has 134 trainable parameters (127 from the ANN, 3 from the *α*-parameters and 4 epidemiological constants *β, σ, γ* and *r*_*d*_) that are calibrated using the data. All the model parameters are calibrated simultaneously. The loss function that we have used is given by:

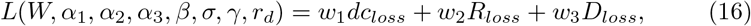

where by *W* we denote the collection of all the ANN weights and biases. The weights *w*_1_, *w*_2_ and *w*_3_ are predefined and they depend on the relative importance of the various data. The terms *dc*_*loss*_, *R*_*loss*_ and *D*_*loss*_ in the above equation denote the loss function contributions coming from the data for daily cases (number of daily infections), total recovered cases and total death cases, respectively for all the days in the calibration period and they are given by the following equations:

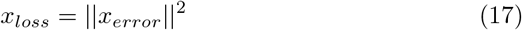

where *x* stands for one of the *dc, R* or *D*, the error terms are defined by the differences of the corresponding model output vectors and the data vectors (with vector components corresponding to the days within the calibration period):

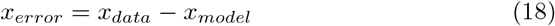

Since we believe that the data is under-reported in general, we assign higher weights to the data points corresponding to the days where the model prediction for any *x* is less than the corresponding data for that *x*. We achieve this by doing the following. After the error vectors are calculated by Eq.(18) we define a new vector 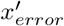 by the following equation:

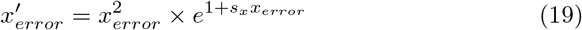

[need to rewrite] Note that in Eq.(19), all operations are performed elementwise. The parameters *s*_*x*_ are pre-defined and depend on *x*. After the vector 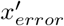 is calculated, we apply historical weighting to the elements of this vector and average them arriving at the norm in Eq.(17):

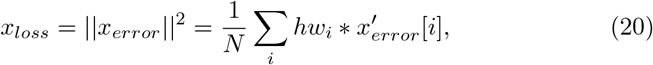

where *N* is the number of days in the calibration period, the sum is over the days in the calibration period and the weight *hw*_*i*_ are predefined historical weights that give more importance to more recent dates in the calibration period.

The algorithm for the model calibration is given in Algorithm 1. Once the model trainable parameters are calibrated, we use these calibrated values to execute the steps in the second loop (i.e., over days in calibration period and future period) of the Algorithm 1 to make future case predictions.

#### Algorithm 1 Model training

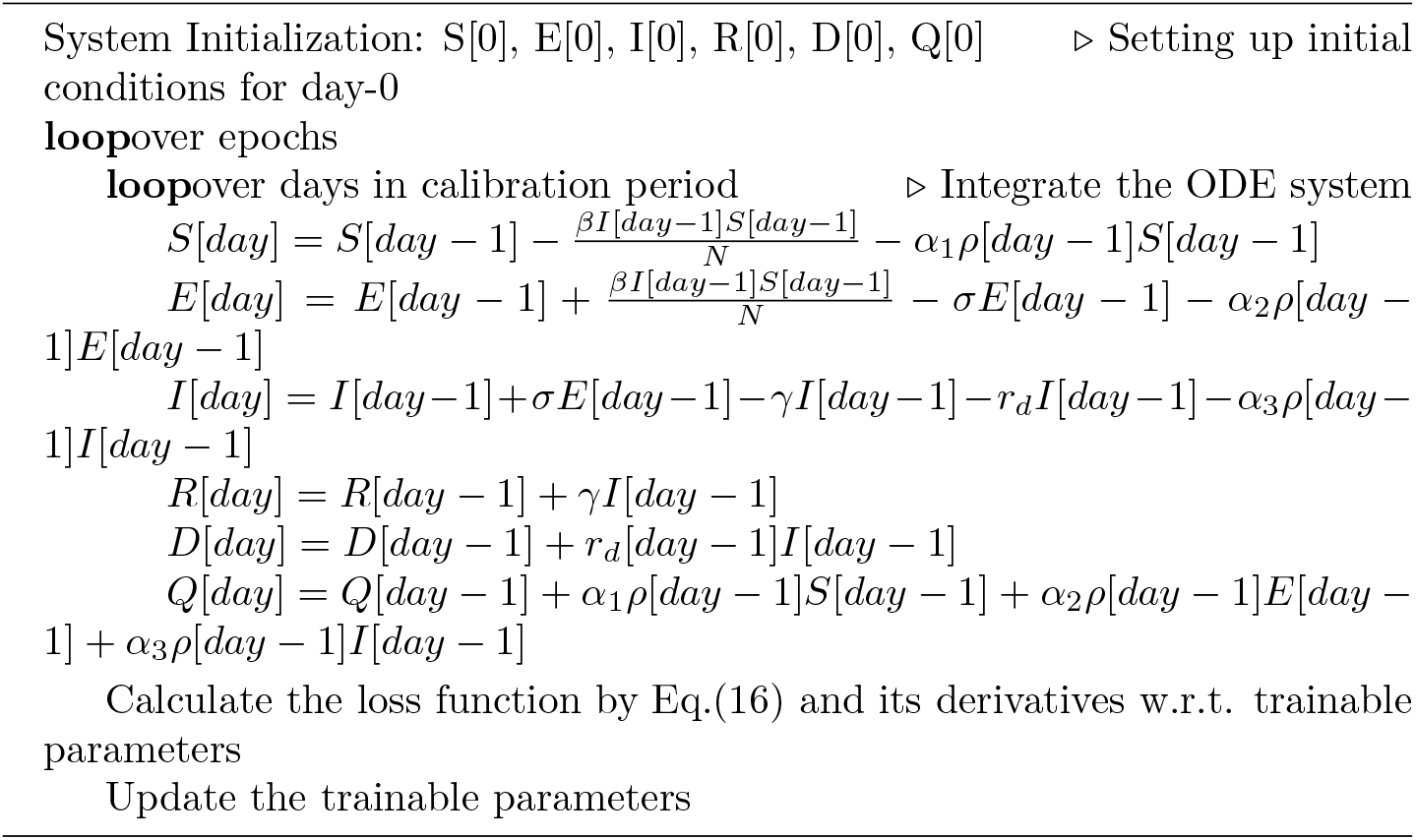

### 4.2 Results

Here we present the model projections with the actual data for US. As it was explained in Section 2.2, the start date of the training period is set to the date when the total confirmed cases are about 500. For the U.S. this corresponds to March 08, 2020 with 518 confirmed cases. For the results presented in this section, the end date of the training period is May 25, 2020. The SEIRDQ model prediction results and the corresponding data for U.S. are presented in Fig.12 and Fig.13 respectively.

**Figure 12:**
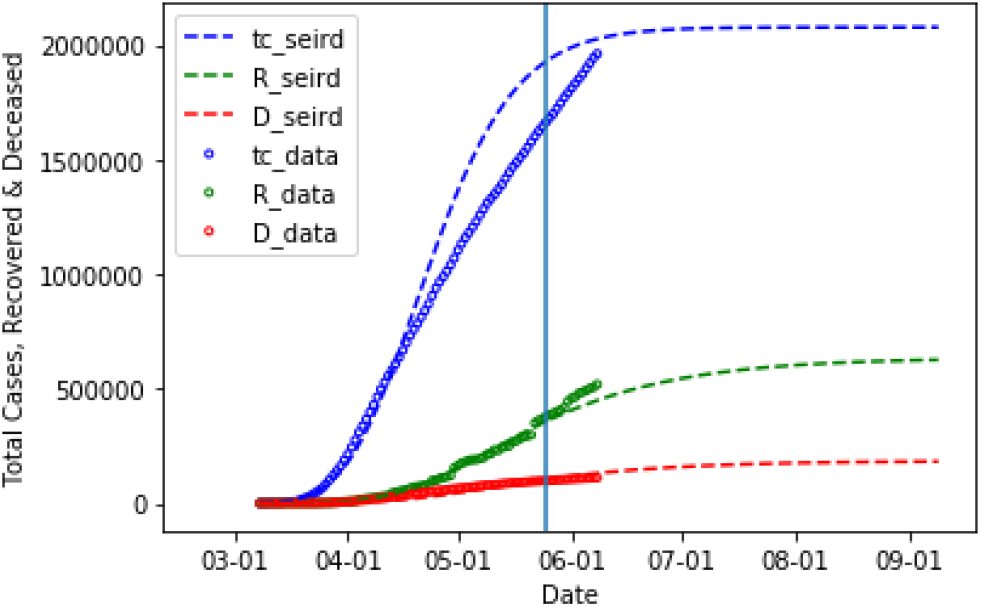
COVID-19 data along with model predictions. “tc” stands for total accumulative cases, “D” stands for total number of deaths and “R” stands for total number of recoveries. The vertical line (on May 25, 2020) separates the training period from prediction period.

**Figure 13:**
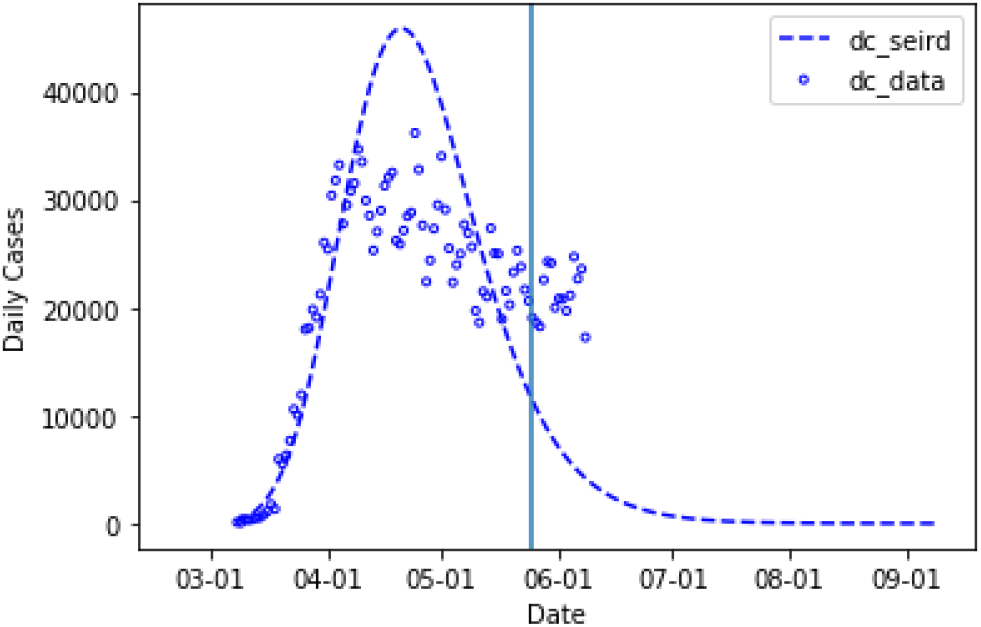
COVID-19 data along with model predictions. “dc” stands for daily cases. The vertical line (on May 25, 2020) separates the training period from prediction period.

### 4.3 Model Benchmarking

In this section, we present model benchmark results where we compare our model predictions for the number of deaths versus that output from other leading models. The benchmark data are sourced from [11] and [12]. We choose the benchmark dates to be June 1, 2020 and June 8, 2020. Training period is from March 8, 2020 through the benchmark date and the projections are made for five days into the future (i.e. until June 6 and June 13). The results are presented in Fig.14.

**Figure 14:**
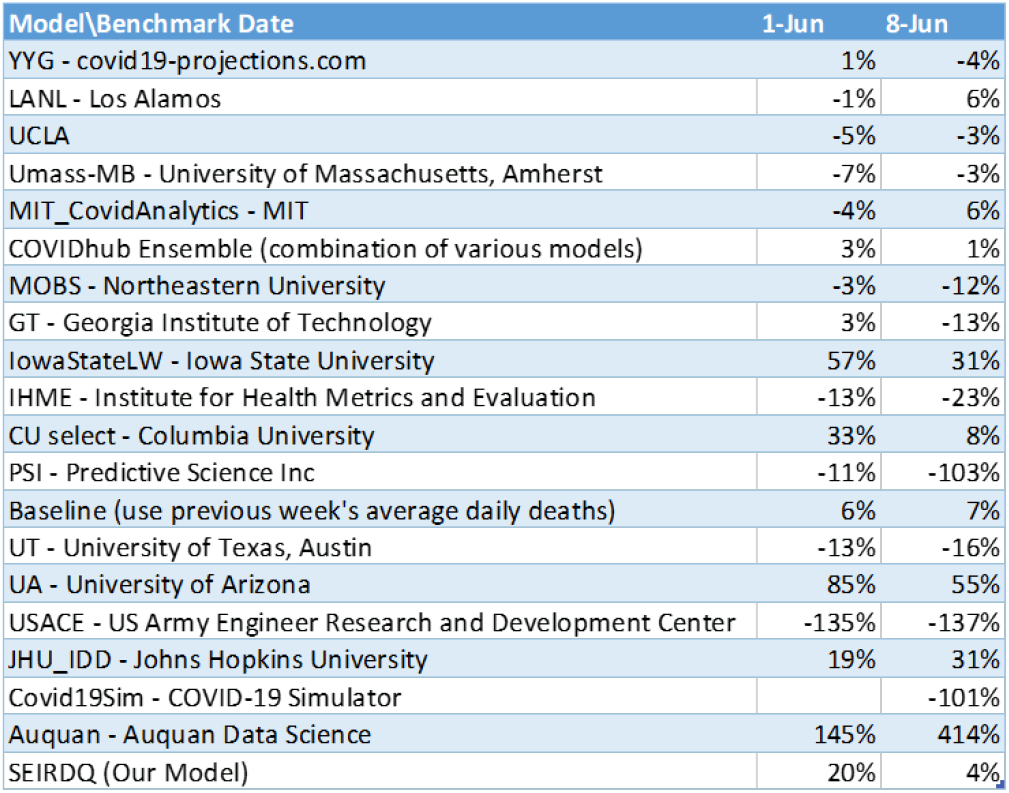
Benchmark against other models. The columns represent percentage difference of model prediction and the actual data for the total number of deaths.

The Fig.14 presents percentage errors of model predicted total number of deaths. Positive error mean over-predicting and negative error means under-predicting.

### 4.4 Modeling Future Waves

Our model has the flexibility to incorporate and predict future waves. This is possible through explicit assumptions on the timings of government directed lock-down and lift-off policies. The way we model the second wave is by turning the quarantine function *ρ* off and on again. We do this by multiplying it with a continuous, turning off and on function that is built using logistic functions:

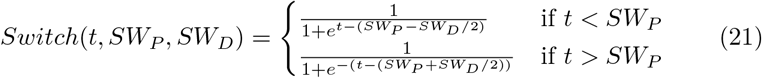

In the above Eq.21 the “Switch”- function depends on two parameters. The first parameter, *SW*_*P*_, is for the second wave position which determines the midpoint of the region where the switch function is zero, and the second parameter, *SW*_*D*_, is for the second wave duration which determines the length of the region where the switch function is zero. As can be seen from the Fig.15, the switch-function smoothly transitions from one to zero then back to one.

**Figure 15.**
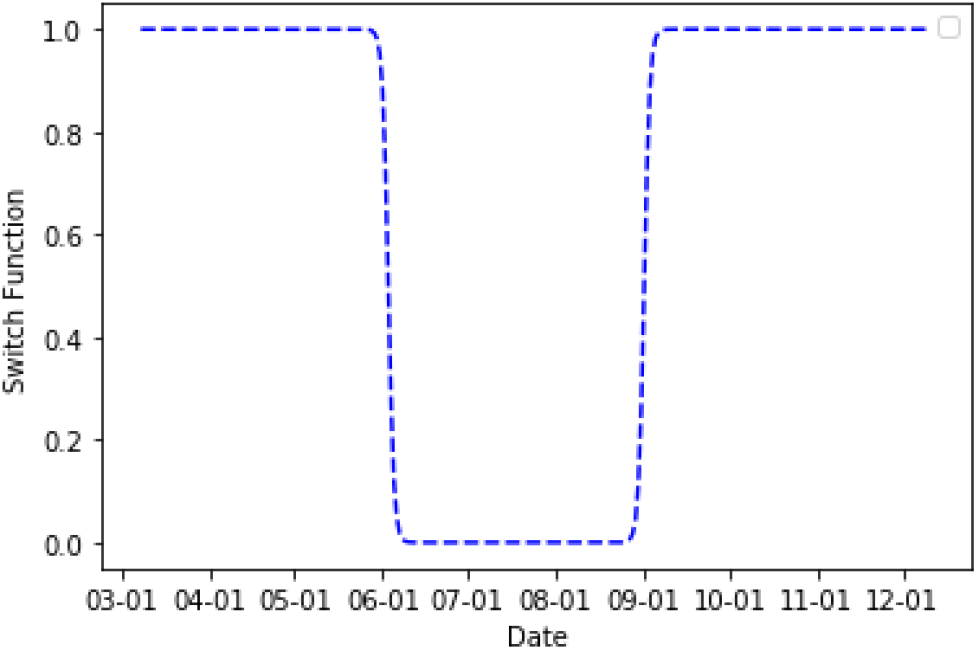

**Figure 16.**
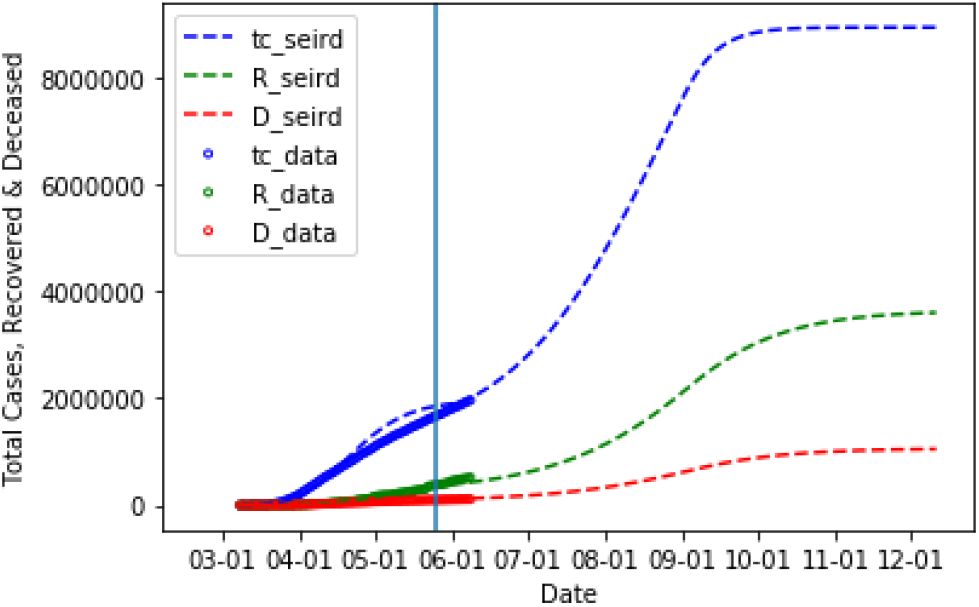

**Figure 17.**
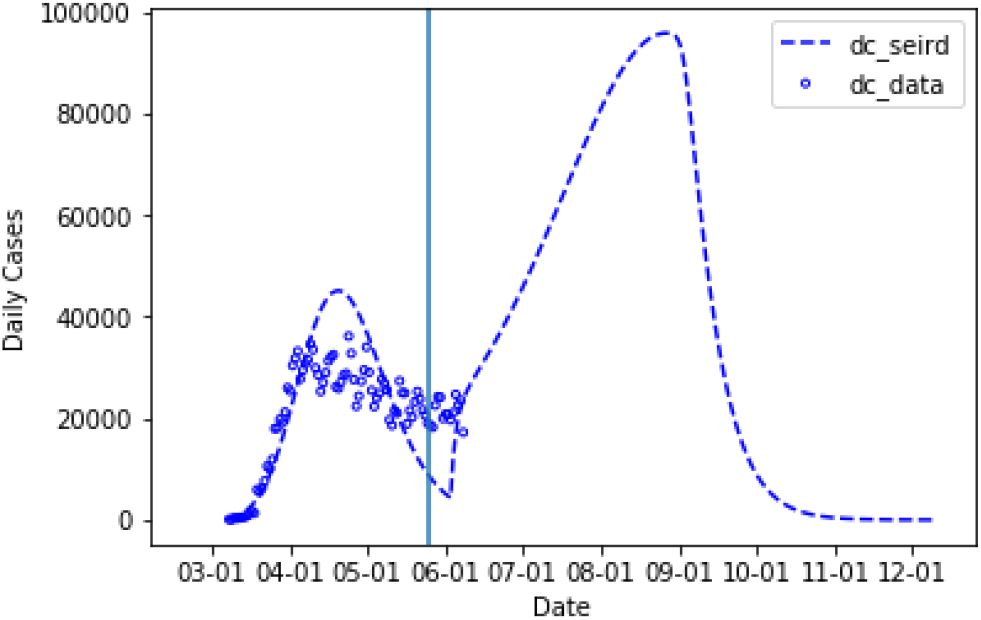

As can be seen from the diagrammatic representation of the model (see Fig.11), people accumulated in the quarantine state - *Q* will not participate in the processes in the top portion of the diagram 11 if we keep the model as is during the second wave. In reality though, as soon as the quarantine policies are lifted, some portion of the people in quarantine state - *Q* that are not infected will become susceptible again (some people start going back to work, schools start resuming classes, etc.). To accommodate this effect we calculate the number of people in the quarantine state - *Q* that moved into that state from the state - *S* until the time when the quarantine policy is lifted. We then take a portion (for the results below, this portion is assumed to be 4%) of that number and move that many people back to the state - *S* on the date when the quarantine policy is lifted.

Assuming the quarantine policies are lifted on July 3, 2020, and they remain open for 90 days we get predictions for the second wave as in the following figures:

## Data Availability

All data produced in the present study are available upon reasonable request to the authors

## Footnotes

1 The worldwide COVID-19 data collected by John Hopkins University is updated daily and available online (https://coronavirus.jhu.edu/map.html)

2 The standard logistic function and its properties are discussed in [13]. For the widely used applications of the S-curve logistic function, the paper [14] provides a scientific basis based on the so called “constructal law” in physics.

